# Predictive modelling of highly multiplexed tumour tissue images by graph neural networks

**DOI:** 10.1101/2021.07.28.21261179

**Authors:** Paula Martin-Gonzalez, Mireia Crispin-Ortuzar, Florian Markowetz

**Affiliations:** Cancer Research UK Cambridge Institute, University of Cambridge, Cambridge, CB2 0RE, UK

**Keywords:** Graph Neural Networks, Highly Multiplexed Imaging, Imaging Mass Cytometry, Tumour Microenvironment, Breast Cancer

## Abstract

The progression and treatment response of cancer largely depends on the complex tissue structure that surrounds cancer cells in a tumour, known as the tumour microenvironment (TME). Recent technical advances have led to the development of highly multiplexed imaging techniques such as Imaging Mass Cytometry (IMC), which capture the complexity of the TME by producing spatial tissue maps of dozens of proteins. Combining these multidimensional cell phenotypes with their spatial organization to predict clinically relevant information is a challenging computational task and so far no method has addressed it directly. Here, we propose and evaluate MULTIPLAI, a novel framework to predict clinical biomarkers from IMC data. The method relies on attention-based graph neural networks (GNNs) that integrate both the phenotypic and spatial dimensions of IMC images. In this proof-of-concept study we used MULTIPLAI to predict oestrogen receptor (ER) status, a key clinical variable for breast cancer patients. We trained different architectures of our framework on 240 samples and benchmarked against graph learning via graph kernels. Propagation Attribute graph kernels achieved a class-balanced accuracy of 66.18% in the development set (N=104) while GNNs achieved a class-balanced accuracy of 90.00% on the same set when using the best combination of graph convolution and pooling layers. We further validated this architecture in internal (N=112) and external test sets from different institutions (N=281 and N=350), demonstrating the generalizability of the method. Our results suggest that MULTIPLAI captures important TME features with clinical importance. This is the first application of GNNs to this type of data and opens up new opportunities for predictive modelling of highly multiplexed images.

## 1 Introduction

Cancer cells in a tumour are embedded in complex tissues including infiltrating immune and inflammatory cells, stromal cells, and blood vessels. The collection of these diverse cells is known as the tumour microenvironment (TME) [24]. Several studies have highlighted the clinical importance of tissue architecture and cell-cell interactions in the TME [12], possibly explained by different selective pressures they exert on tumour evolution [22]. Capturing spatial organization of the TME and its impact on patients is thus a critical problem in cancer medicine.

The study of the TME has been boosted by the development of highly multiplexed imaging technologies, which measure a large number of markers per tissue section and capture both the complex cellular phenotypes in the TME and their spatial relationships [7]. One example of highly multiplexed imaging is Imaging Mass Cytometry (IMC), which enables imaging at cellular resolution by labelling tissue sections with metal-isotope-tagged antibodies [5] (Figure 1a). These novel imaging technologies have resulted in many data analysis challenges, including the segmentation of cell nuclei and quantification of proteins [21,20] and the phenotypic clustering of cells [1]. IMC technologies are commercially available and start being used routinely world-wide, which increases the amount of available data, but also its technical variability, and new strategies for data standardization will be needed.

**Fig. 1.**
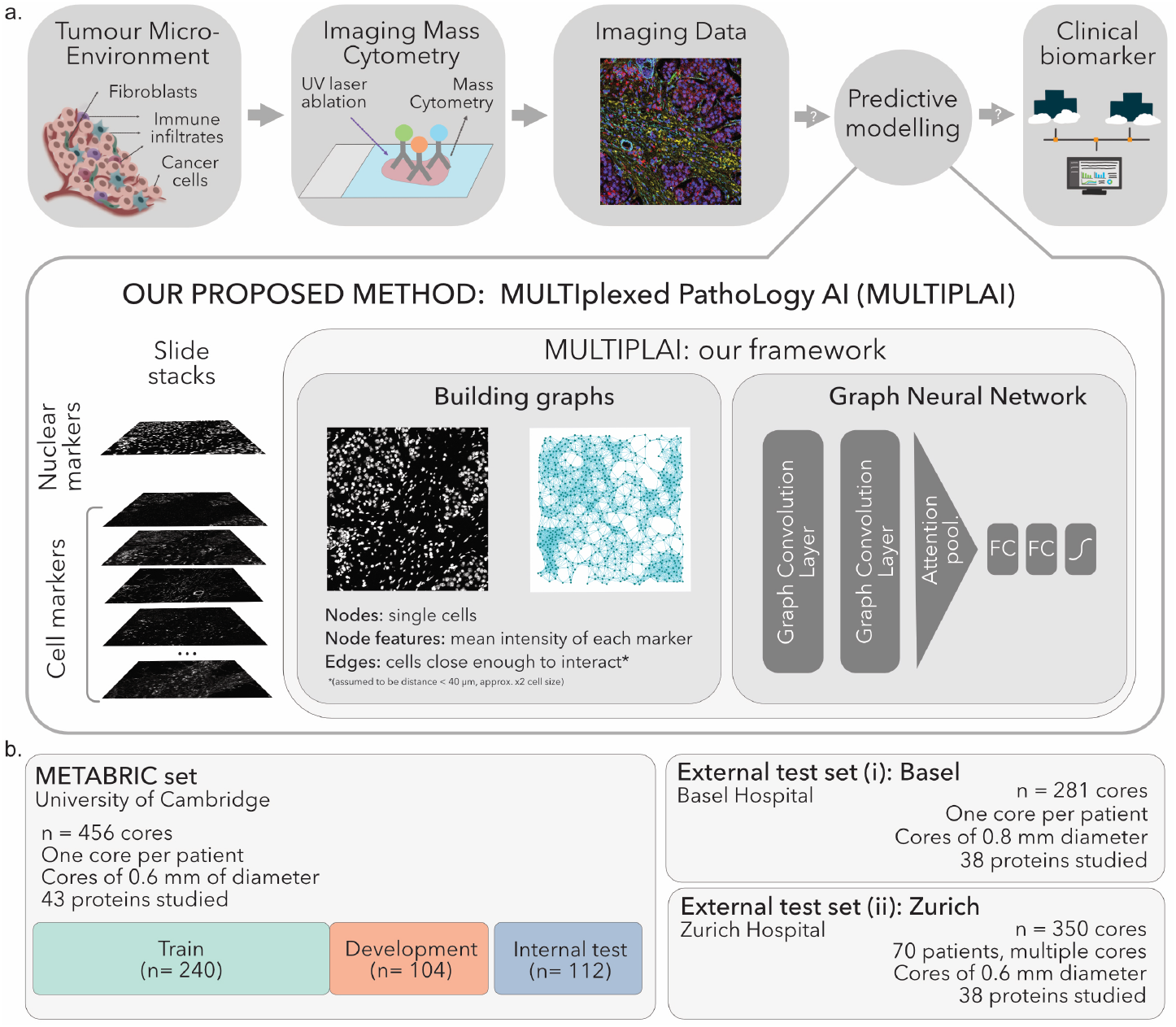
(a) Overview of the proposed method. MULTIPLAI takes IMC imaging data as input for a GNN model to predict clinical biomarkers (b) Training and independent test sets used for the analysis.

A natural approach to rigorously capture the information provided by IMC is graph representation learning. Graph kernels have been developed for similar problems [15,2], and recent developments in Geometric Deep Learning via Graph Neural Networks (GNNs) have shown how to analyse interactions and topology in a data-driven manner [3,4].

Here we propose using graph representation learning via GNNs to study IMC phenotypes together with spatial cell-to-cell interactions. We describe and evaluate MULTIPLAI (MULTIplexed PathoLogy AI), a framework that applies GNNs on highly multiplexed images for data-driven exploration of patterns in the TME. In this proof-of-concept study we used MULTIPLAI to predict Oestrogen Receptor (ER) status, a key biomarker that determines treatment options for breast cancer patients [18]. ER-positive and ER-negative breast cancers are known to be different at the histopathological, molecular, and clinical level [6,19], which makes ER-subtyping an excellent first case study to explore and benchmark the ability of MULTIPLAI to simultaneously capture the phenotypic and spatial structure of the tumour tissue. To evaluate our method, we trained different architectures for MULTIPLAI (Figure 1a) and benchmarked them against the Propagation Attribute Graph Kernel [17]. Then, we applied the best MULTIPLAI model to internal and external test sets (Figure 1b) and compared the effects of different data standardization strategies.

To our knowledge, MULTIPLAI is the first framework that applies GNNs to highly multiplexed tumour tissue images. The method captures spatial interactions between complex cell phenotypes and their relationship with clinically-relevant biomarkers, opening the door to a new understanding of the dynamics and mechanisms of the TME.

## 2 Method

MULTIPLAI combines two key steps: the construction of a graph from imaging data, and the application of GNNs to predict clinical variables from these graphs.

### 2.1 Building graphs from highly multiplexed imaging

We represent an IMC image as an undirected graph *G* = (*V, E*) consisting of sets of nodes *V* and edges *E* ⊆ *V* ×*V*. Each node corresponds to a feature vector *x*_*v*_ ∈ *R*^*m*^ where *m* is the number of features. The data sets we used were already segmented and we defined each cell to be a node in *V*. We defined the node features *x*_*v*_ as the mean expression of each protein analysed. The edge set *E* is built by connecting cells whose segmentation centroids are within 40 *µ*m from each other. This number is approximately twice the size of a regular cell [8] as we assume that cells have to be within reach to each other to interact [12].

### 2.2 Graph Neural Networks

GNNs are a type of neural network operating on a graph structure. They use an iterative procedure based on message passing from nodes to their neighbours [25]. GNNs used for graph-level binary classification contain two characteristic building blocks, graph convolution layers and graph pooling layers, that can also be combined with fully connected layers. The MULTIPLAI framework consists of the following graph convolution and pooling layers:

#### Graph convolution layers

Graph convolution layers are where the iterative procedure takes place [13]. The goal is to build an encoder that maps nodes to *d*-dimensional embeddings. Such embeddings are built using message propagation in local neighborhoods. On each hidden layer, message passing takes place and each node feature vector is updated considering information from its neighbours:

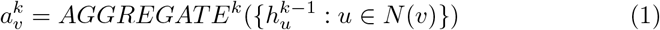

*and*

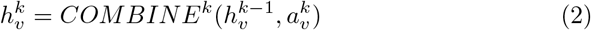

where *N* (*v*) is the neighborhood of each node *v*, 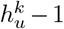 represents the feature vector of each node in layer *k* − 1 and 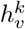 refers to the updated feature vector for node *v* after layer *k*.

#### Graph pooling layers

After the neighborhood information has been propagated through the graph, the resulting node embeddings need to be combined to generate graph-level features *h*_*g*_ necessary for graph classification. For that, the updated node feature vectors have to be combined as follows:

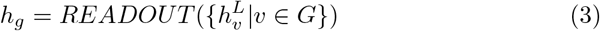

Where 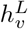 is the updated feature vector of each node *v* after the last layer *L*. We can combine node features using the average or the sum of the features as a readout. Another way to combine updated node features is to incorporate attention. Attention mechanisms [23] allow the network to focus on parts that are more relevant to the task being learnt. In the context of graph pooling, we can allow the network to decide the amount of focus given to each of the nodes in the graph as proposed in [16]:

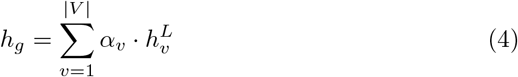

where *h*_*g*_ is the graph-level feature vector resulting from the combination of updated node features, |*V*| is the number of nodes in the graph and *α*_*v*_ weights the attention given to each node.

Attention is calculated by feeding the updated node vectors into a parallel neural network to learn the weight given to each node through gradient descent. Thus, each *α*_*v*_ from Equation 4 is calculated as:

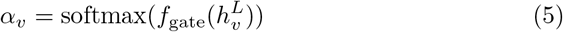

where the softmax function ensures *α*_*v*_ ∈ (0, 1) and ∑_*V*_ *α*_*v*_ = 1, *f*_gate_ is the neural network used for learning the attentions and 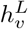 are the node vectors.

#### Benchmarking

There are no state of the art methods are available to perform spatial analysis of highly multiplexed tissue data in a data driven manner to benchmark the performance of GNNs against. Therefore, we will use as a baseline the graph representation approach that uses previously defined Graph Kernels handling graphs with continuous node features. In particular, we will use the Propagation Attribute Kernel [17].

## 3 Data sets

The data we used for the train, development and internal test sets is publicly available and has been previously described [1]. Briefly, this cohort consisted of 456 patients diagnosed with primary invasive breast carcinoma in Cambridge, UK. Cores of 0.6 mm in diameter were acquired and tissue micro-arrays were constructed and stained with a panel of 43 antibodies conjugated with metal tags. The slides were then analysed using the Hyperion Imaging Mass Cytometer (Fluidigm). We used the cell segmentations and marker quantification provided by the original publication [1]. The clinical information of these patients, including ER status, is also publicly available. We randomly split these cores into three data sets: training (n = 240), development (n = 104) and internal test sets (n = 112).

The data we used as the external test sets is also publicly available [9]. The first external test set contained 281 patients from the University Hospital of Basel. Pathologists reported clinical information and supervised the tissue microarray construction of cores of 0.8 mm. The second external test set contained 70 patients from University Hospital of Zurich Hospital. Patients had multiple cores of 0.6 mm acquired and the data set contains a total of 350 slides. We used all the slides with their patient’s ER status label. In both cases, samples were stained with a panel of 38 antibodies coupled to metal tags. An Hyperion Imaging System (Fluidigm) was used for acquisition. The segmentation of cells and quantification of markers again were provided by the original publication [9]. Clinical features of all cohorts are compared in Supplementary Figure 1.

## 4 Experiments and Results

The aim of our proof-of-concept study is to evaluate the potential of Graph Representation learning anf GNNs on IMC. Here, we focus on the well established and clinically important TME differences between ER-positive (ER+) and ER-negative (ER-) breast cancers and study how well it can be captured by MULTIPLAI, our proposed GNN-based framework. The code can be accessed here: https://github.com/markowetzlab/MULTIPLAI

### 4.1 Marker selection and preprocessing

We only considered markers present in all the three cohorts (Metabric, Basel and Zurich). Since our goal is to quantify differences in the TME between ER+ and ER-patients, we excluded the ER marker itself as well as all other markers with highly correlated expression at the cellular level. In this way, our model focusses on the cellular composition and spatial distribution of the microenvironment and is not confounded by individual proteins. Requiring a Spearman *ρ* greater than 0.65 and a p-value smaller than 0.05 after Bonferroni correction resulted in the exclusion of HER2 (*ρ* = 0.670), Progesterone Receptor (*ρ* = 0.739), GATA3 (*ρ* = 0.803), E-Cadherin (*ρ* = 0.699) and PARP/Casp 3 (*ρ* = 0.671). After the correlation removal step we ended up with a set of 19 proteins from the immune system, extracellular matrix, cytokeratins and cell processes to be used (Supplementary Materials, Table 1). We used the logarithmic transform of intensities for the analysis, setting null intensities to 10^*−*9^. We used the training set to define the standard scaling parameters for each marker in the single cells, and scaled the development set accordingly. In addition, we evaluated different scaling strategies to account for the different image acquisition protocols followed in each of the test datasets (Section 4.3).

**Table 1.** Models tried for predicting ER status from IMC images. PA stands for Propagation Attribute. GNN stands for Graph Neural Network. Binary Cross Entropy (BCE) loss and Class Balanced (CB) accuracy reported are from the development set.

| Model | Architecture |  | Best model |  |
| --- | --- | --- | --- | --- |
|  | Number of layers | Pooling | BCE Loss | CB Accuracy |
| PA Graph Kernel | N.A. |  | 7.306 | 66.18 |
| GNN | 1 | Mean | 0.174 | 78.4 |
|  |  | Sum | 0.353 | 85.72 |
|  |  | Attention | 0.199 | 86.86 |
|  | 2 | Mean | 0.162 | 86.00 |
|  |  | Sum | 0.278 | 81.16 |
|  |  | **Attention** | **0.159** | **90.00** |

### 4.2 Finding the best architecture

As a baseline method to check the potential of graph representation in this problem, we decided to use the Propagation Attribute graph kernel. This model was implemented in Grakel (version 0.1b7) and sklearn (version 0.22.1). The preserved distance metric on local sensitive hashing was L1 normalization. We performed a grid search of the bin width parameter (1,3,7,9 and 15) selecting the model that resulted in lower BCE loss.

For MULTIPLAI, we designed six GNN architectures with different numbers of graph convolution layers and different pooling methods (section 2.2). A graphical description is shown in Supplementary Figure 2. A single fully connected layer served as *f*_*gate*_ for the attention pooling. Graph level features obtained after pooling in all cases were fed into two fully connected layers and a sigmoid function, whose outputs were rounded to obtain binary predictions. We used Binary Cross Entropy (BCE) Loss and Adam optimizer. Architectures were implemented using Deep Graph Library (version 0.4.3) and Pytorch (version 1.5.1).

We trained each architecture on the training set using mini-batches, and for each epoch we evaluated the loss and class-balanced accuracy in the development set. We implemented early stopping by finding the minimum development set loss and allowing 20 epochs of patience before stopping. We performed a grid search for hyper-parameter optimization: batch size of 10, 20 and 30, hidden dimensions of fully connected layers 50, 100 and 150, dropout of 0.3 and 0.5, learning rate of 0.01 and 0.001, weight decay of 0.1, 0.01 and 0.01 and graph hidden dimensions of 50, 100 and 150 in the cases with two graph convolutions. In each case, we optimised each hyperparameter to give the a lower loss and lower standard deviation in the development set (Table 1).

We found some signal showing in the graph kernel model (Class Balanced Accuracy of 66.18) and, when moving to GNNs, even shallow architectures were able to capture the TME differences in ER+ and ER-patients with the lowest balanced accuracy being 78.40. This suggests the potential of graph representation learning for this problem and overall, GNN models do significantly better than the Graph Kernel. Additionally, the losses in the models with two layers (0.162, 0.278 and 0.159) are significantly lower than those with only one (0.174, 0.353 and 0.199). We selected as the best model the combination of two graph convolutions and attention pooling for the next sections as it reached the lower binary cross entropy loss (0.159) on the development set.

### 4.3 Comparing standardization strategies

We then applied the best model identified in the last section to the internal and two external test cohorts (Figure 1b). This step is complicated by systematic biases in the data: The three cohorts come from three different institutions, use different sizes of tissue, use different numbers of markers and show differences in the global staining distribution across slides. To alleviate these biases, we compared two different stain standardization strategies.

#### Strategy 1: fixed scaling

The best architecture identified in section 4.2 was applied to the internal and external test cohorts (Table 2, Supplementary table 2), using the same fixed standard scaling applied on the development set. A balanced accuracy of 78.96 is seen in the internal test cohort but performance declines in the Zurich and particularly the Basel set (balanced accuracies of 68.19 and 60.16 respectively). An analysis of the stain histograms (Supplementary materials, figure 3a) reveals that the two external sets are systematically different. Since IMC has been used so far only on same-institution descriptive studies, the field has not yet come across the need to define standardization strategies to enable inter-institution comparisons. To correct for the biases, we compared an additional normalization strategies.

#### Strategy 2: independent scaling

To explore whether we could correct for the staining differences, we scaled the intensity distributions for each marker on each dataset independently (Supplementary materials, figure 3b). When validating the initial model from section 4.3 using this version of scaling, the performance on the external test sets improves significantly (from a balanced accuracy of 60.16 to 65.71 in the Basel set and from 68.19 to 75.10 in the Zurich set) (Table 2, Supplementary table 2). Importantly, the performance degradation observed in the external sets in all strategies reflected known experimental differences. The Basel dataset, in particular, had a bigger core size (0.8 mm) than the rest of the sets (0.6 mm), resulting in significantly larger graphs than the other two.

**Table 2.** Metrics for each of the test cohorts and preprocessing conditions

|  | Class Balanced Accuracy |  |
| --- | --- | --- |
|  | Strategy 1 | Strategy 2 |
| Internal test (Metabric) | 78.96 | **78.96** |
| External test (Basel) | 60.16 | **65.71** |
| External test (Zurich) | 68.19 | **75.10** |

## 5 Discussion

We have presented a proof-of-concept study showing how GNNs can be applied to IMC data for predictive modelling of clinical biomarkers. We showed that our proposed framework, called MULTIPLAI, was able to capture TME differences between ER+ and ER-patients. These results signal a promising new avenue for the computational analysis of highly multiplexed images.

Some features of the method can be further optimized: First, we pragmatically fixed the distance threshold for building the graph and we plan to perform an ablation study of this distance to check the stability of the graph construction step.

Second, the influence of scaling and graph size to model performance suggests that IMC data standarization is a key challenge to develop methods that are robust across institutions. Data augmentation could help increasing the robustness of the MULTIPLAI framework to these changes between institutions but there is little literature available on data augmentation for graphs [26,14]. This suggests that the need to propose such methods is also an open challenge in the GNN field. The descriptive work we presented in Section 4.2 could be used as a benchmark for new data augmentation techniques.

Third, it will be important to address the explainability of the framework in future work. Exploring the node and feature importance driving different predictions will help cancer biologists understand that the method is capturing known TME differences as well as discover new patterns not yet reported. Unlike in the Convolutional Neural Networks domain, very few alternatives are available to study the explainability of decisions in GNNs [11,10], and more work in the area is needed.

Finally, we compared the suitability of two graph learning approaches (GNNs and Graph Kernels) in this study but it would be interesting to compare the performance of graph learning to Convolutional Neural Networks (CNNs) in this setting. Although off-the-shelf CNNs are built for RGB images, the input tensor could be extended to match the number of channels in IMC images.

To conclude, this proof-of-concept study shows that the analysis of highly multiplexed images in cancer can benefit from using GNN-based methods. These frameworks spatially integrate the different phenotypes of the TME that could be used to perform data-driven clinical decisions.

## Data Availability

The data used is publicly available in their respective publications. The framework we developed is publicly available here: https://github.com/markowetzlab/MULTIPLAI

https://www.nature.com/articles/s43018-020-0026-6

https://www.nature.com/articles/s41586-019-1876-x

https://github.com/markowetzlab/MULTIPLAI

