## Supplementary materials for "Predictive modelling of highly multiplexed tumour tissue images by graph neural networks"

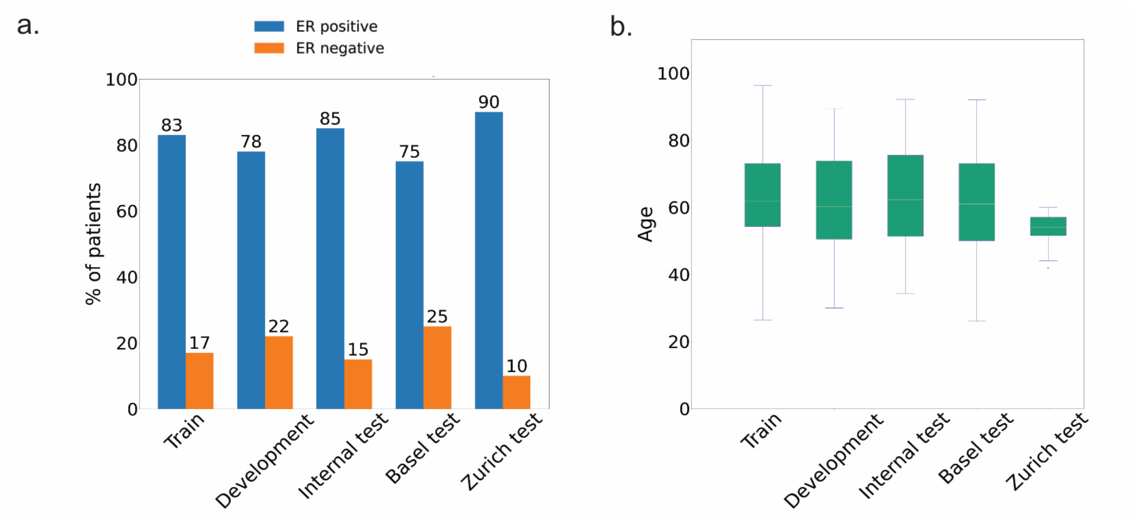


**Fig. 1.** (a) Proportion of ER+ and ER- patients in each cohort. (b) Age distribution in each cohort

**Table 1.** Markers used for the analysis and their function in breast cancer and tumour biology

| **Immune cells** | |
|---|---|
| CD68 | Macrophages |
| CD3 | T-cells |
| CD45 | Memory T-cells |
| CD20 | B-cells |
| CD31 | T-cell activation |

| **Cell processes** | |
|---|---|
| TP53 | Regulates cell cycle |
| Ki67 | Proliferation marker |
| CD44 | Cancer stem cells |
| PS6 | Part of MAPK pathways |
| EGFR | Involved in cancer pathogenesis |

| **Cytokeratins** | |
|---|---|
| CK19 | Luminal epithelial cell |
| CK8/18 | Epithelial intermediate filament |
| CK14 | Basal-like breast cancer |
| CK7 | Hallmark for breast cancers |
| PanCK | Epithelial tumour |
| CK5 | Epithelial cell layer |

| **Extracellular matrix** | |
|---|---|
| Vimentin | Intermediate filament |
| Fibronectin | Extracellular matrix |
| SMA | Smooth Muscle Actin |

**Table 2.** Elaborated metrics from the different validation set and each scaling strategy

| | | Internal test (Metabric) | External test (Basel) | External test (Zurich) |
|---|---|---|---|---|
| Strategy 1 | Balanced Accuracy | 78.96 | 60.16 | 68.19 |
| | Recall | 96.8 | 94.28 | 93.51 |
| | Precision | 92.86 | 78.81 | 93.81 |
| Strategy 2 | Balanced Accuracy | 78.96 | 65.71 | 75.10 |
| | Recall | 96.8 | 81.43 | 87.35 |
| | Precision | 92.86 | 82.61 | 95.61 |

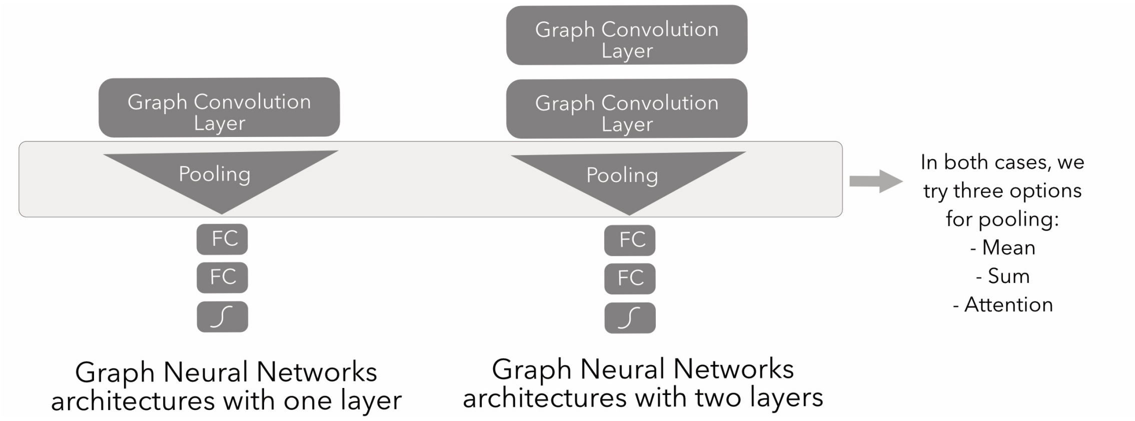


**Fig. 2.** Summary details of the graph neural network architectures studied.

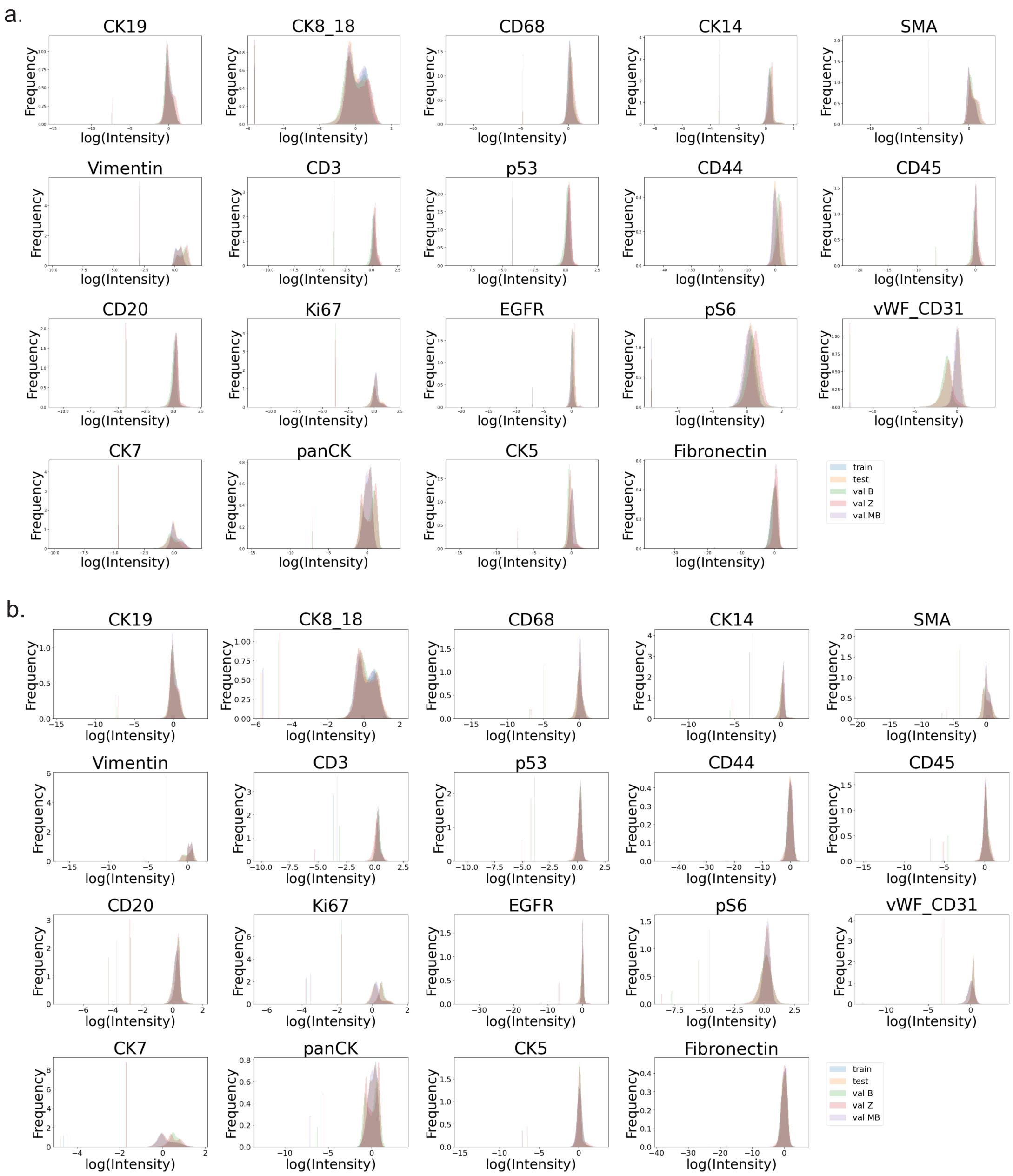


**Fig. 3.** (a) Histogram of log (intensities) for each marker and cohort scaled using standard scaling with fixed parameters from the train set. (b) Histogram of log (intensities) for each marker and cohort scaled using standard scaling with parameters set from each cohort independently.
